# Impact of COVID-19 shelter-in-place order on transmission of gastrointestinal pathogens in Northern California

**DOI:** 10.1101/2021.01.12.21249708

**Authors:** Philip L. Bulterys, Nicole Y. Leung, Atif Saleem, Indre Budvytiene, Niaz Banaei

**Affiliations:** Department of Pathology, Stanford University School of Medicine, Stanford, CA, USA; Department of Psychiatry and Behavioral Sciences, Stanford University School of Medicine, Stanford, CA, USA; Department of Medicine, Division of Infectious Diseases and Geographic Medicine, Stanford University, Stanford, CA, USA; Clinical Microbiology Laboratory, Stanford University Medical Center, Stanford, CA, USA

**Keywords:** Gastrointestinal pathogen, gastroenteritis, COVID-19, shelter-in-place, community transmission

## Abstract

Society-wide cessation of human interaction outside the household due to the COVID-19 shelter-in-place created a unique opportunity in modern history to reexamine the transmission of communicable gastrointestinal pathogens. We conducted a quasi-experimental study from January 1, 2018 to Sept 30, 2020 to investigate the effect of California’s COVID-19 shelter-in-place order on the community transmission of viral, bacterial, and parasitic gastrointestinal pathogens detected with the FilmArray GI Panel (BioFire Diagnostics, Salt Lake City, UT). The incidence of viral causes of gastroenteritis, enteroaggregative/enteropathogenic/enterotoxigenic *Escherichia coli, Shigella*, and C*yclospora cayetanensis* decreased sharply after shelter-in place took effsect, whilst *Salmonella, Campylobacter*, shiga toxin-producing *E. coli* (O157 and non-O157) and other bacterial and parasitic causes of gastroenteritis were largely unaffected. Findings suggest community spread of viral gastroenteritis, pathogenic *E. coli* (except for shiga toxin-producing *E. coli*), *Shigella*, and *Cyclospora* is more susceptible to changes associated with shelter-in-place than other gastrointestinal pathogens.

## Brief Report

In response to the COVID-19 pandemic, California was the first state to impose a strict shelter-in-place (SIP) order in March 2020 (1). Although enforced social distancing early in the pandemic appears to have delayed the spread of COVID-19 (1, 2), little is known about its potential impact on the incidence of other communicable infectious diseases. Such a natural experiment involving the society-wide cessation of human interaction outside the household is unique in modern history and could provide useful insight regarding transmission patterns of other pathogens circulating in the community. The objective of this study was to determine the impact of California’s SIP order on the gastrointestinal pathogen landscape dynamics in Northern California.

The FilmArray GI Panel (BioFire Diagnostics, Salt Lake City, UT) is a multiplex, on-demand, sample-to-answer, real-time PCR assay for the syndromic diagnosis of infectious gastroenteritis (3). We analyzed all FilmArray GI Panel results for adult and pediatric patients at the Stanford Health Care Clinical Microbiology laboratory between January 1, 2018 and September 30, 2020. For inpatients, rejection criteria included excluding patients that had been hospitalized for more than 72 hours. We calculated odds ratios and performed Fisher’s exact tests to compare the test positivity rates of all 22 pathogens represented on the panel before and after March 20, 2020, when California’s SIP order was instituted. The study was approved by the Stanford Institutional Review Board.

In total, 10,317 tests were performed during the study period (from 8,321 unique patients) and were included in the analysis. Of these, 8,677 were performed before SIP and 1,640 were performed after SIP. The average number of tests performed per quarter for the entire study period (January 1, 2018-September 30, 2020) was 938 (see Figure 1). Of all tests performed before SIP (n = 8,677), 4,792 (55%) were from patients admitted to a Stanford-affiliated hospital, and 3,885 (45%) were from patients seen in the outpatient setting. Of all tests performed after SIP (n = 1,640), 1,045 (64%) were from inpatients, and 595 (36%) from outpatients.

**Figure 1.**
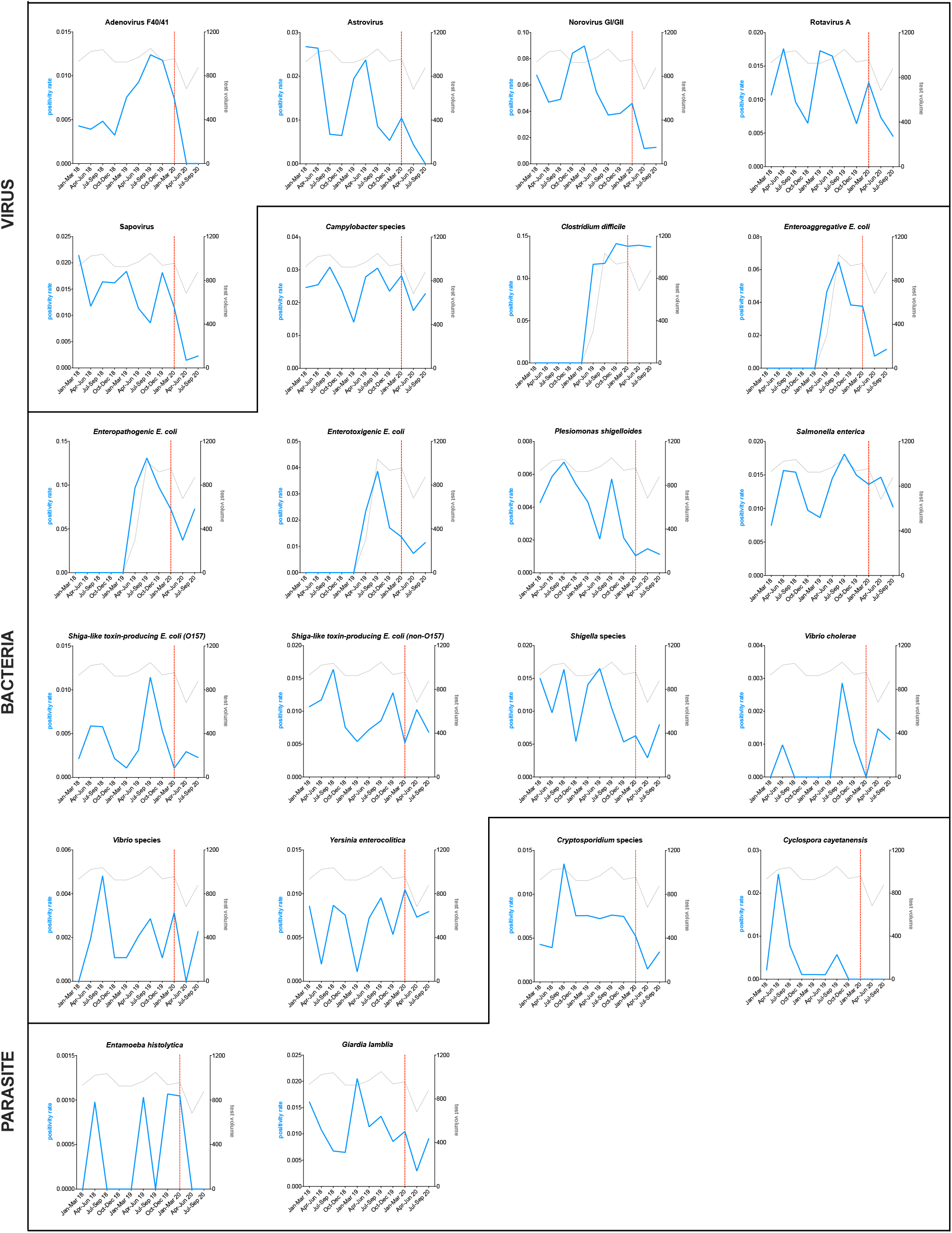
GI PCR positivity rates before and after implementation of California’s shelter-in-place, March 20, 2020. Shown are the quarterly positivity rate (positives/tests) for all gastrointestinal pathogens represented on the FilmArray GI panel from January 1, 2018 to September 30, 2020. The blue line in each plot indicates test positivity rate, whereas the grey line indicates test volume (total tests per quarter). The dotted red line designates when California’s shelter-in-place was instituted (March 20, 2020). The plots are organized with viruses appearing first, then bacteria, then parasites.

When comparing rates of infection before SIP (January 1, 2018-March 19, 2020) to after SIP (March 20, 2020-September 30, 2020), we found that the incidence of viral causes of gastroenteritis, including adenovirus F40/41, astrovirus, norovirus, rotavirus, and sapovirus, was markedly decreased after shelter-in place took effect (see Figure 1). Adenovirus F40/41 detection decreased by a factor of 13 (p = 0.0004), astrovirus by a factor of eight (p < 0.0001), norovirus by a factor of five (p < 0.0001), rotavirus by a factor of two (p = 0.03), and sapovirus by a factor of eight (p < 0.0001) (see Table 1). All pathogenic *Escherichia coli* types (with the exception of shiga toxin-producing *E. coli*), *Shigella* species, and C*yclospora cayetanensis* were also detected significantly less frequently after shelter-in-place. The incidence of enteroaggregative *E. coli* decreased by a factor of four (p < 0.0001), enteropathogenic *E. coli* by a factor of two (p < 0.0001), enterotoxigenic *E. coli* by a factor of three (p < 0.0001), *Shigella* species by a factor of two (p = 0.033), and C*yclospora cayetanensis* positivity rates decreased from 0.51% to 0% (p = 0.0019). The incidence of *Plesiomonas shigelloides, Cryptosporidium* species, *Entamoeba histolytica*, and *Giardia lamblia* also decreased, but the changes were not statistically significant. The incidence of other gastrointestinal pathogens, including *Campylobacter* species, *Clostridium difficile, Salmonella enterica*, shiga toxin-producing *E. coli* (O157 and non-O157), *Vibrio cholerae* and other *Vibrio* species, and *Yersinia enterocolitica*, was largely unaffected.

**Table 1.**
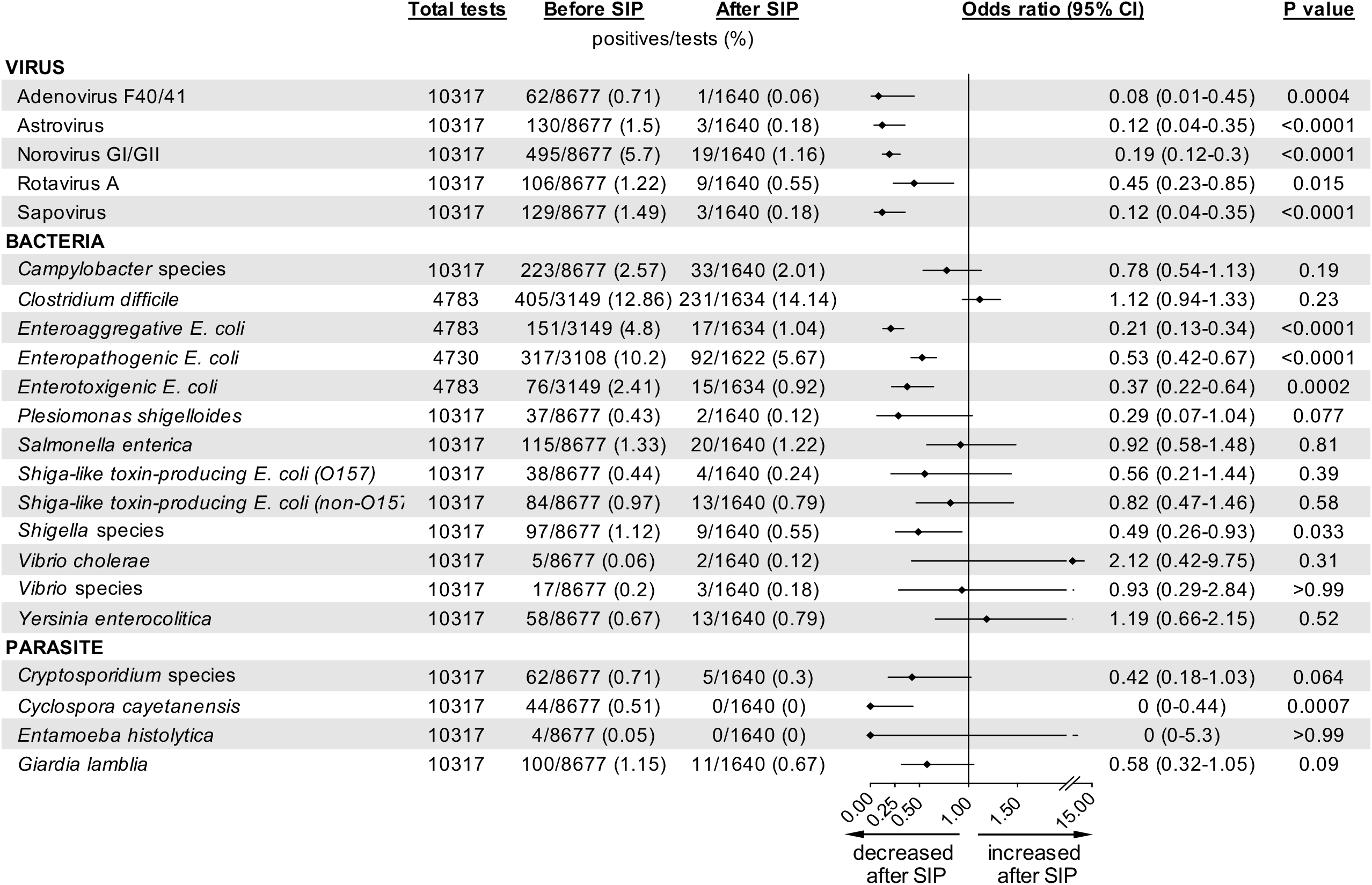
The odds of detection of gastrointestinal pathogens before and after implementation of California’s shelter-in-place, March 20, 2020. Shown for each pathogen on the FilmArray GI panel are the total number of tests performed, the test positivity rates before and after shelter-in-place was instituted, the odds ratio and 95% confidence interval (CI) comparing the odds of test positivity before SIP/after SIP, and P value calculated using a Fisher’s exact test. Odds ratios were calculated using the Baptista Pike method. All statistical comparisons were implemented in GraphPad Prism version 9.0.0.

|  | <u>Total tests</u> | <u>Before SIP</u> | <u>After SIP</u> |  | <u>Odds ratio (95% CI)</u> | <u>P value</u> |
| --- | --- | --- | --- | --- | --- | --- |
|  |  | positives/tests (%) |  |  |  |  |
| VIRUS |  |  |  |  |  |  |
| Adenovirus F40/41 | 10317 | 62/8677 (0.71) | 1/1640 (0.06) | ◆— | 0.08 (0.01-0.45) | 0.0004 |
| Astrovirus | 10317 | 130/8677 (1.5) | 3/1640 (0.18) | ◆— | 0.12 (0.04-0.35) | <0.0001 |
| Norovirus GI/GII | 10317 | 495/8677 (5.7) | 19/1640 (1.16) | ◆— | 0.19 (0.12-0.3) | <0.0001 |
| Rotavirus A | 10317 | 106/8677 (1.22) | 9/1640 (0.55) | ◆— | 0.45 (0.23-0.85) | 0.015 |
| Sapovirus | 10317 | 129/8677 (1.49) | 3/1640 (0.18) | ◆— | 0.12 (0.04-0.35) | <0.0001 |
| BACTERIA |  |  |  |  |  |  |
| *Campylobacter* species | 10317 | 223/8677 (2.57) | 33/1640 (2.01) | ◆— | 0.78 (0.54-1.13) | 0.19 |
| *Clostridium difficile* | 4783 | 405/3149 (12.86) | 231/1634 (14.14) | ◆— | 1.12 (0.94-1.33) | 0.23 |
| *Enteroaggregative E. coli* | 4783 | 151/3149 (4.8) | 17/1634 (1.04) | ◆— | 0.21 (0.13-0.34) | <0.0001 |
| *Enteropathogenic E. coli* | 4730 | 317/3108 (10.2) | 92/1622 (5.67) | ◆— | 0.53 (0.42-0.67) | <0.0001 |
| *Enterotoxigenic E. coli* | 4783 | 76/3149 (2.41) | 15/1634 (0.92) | ◆— | 0.37 (0.22-0.64) | 0.0002 |
| *Plesiomonas shigelloides* | 10317 | 37/8677 (0.43) | 2/1640 (0.12) | ◆— | 0.29 (0.07-1.04) | 0.077 |
| *Salmonella enterica* | 10317 | 115/8677 (1.33) | 20/1640 (1.22) | ◆— | 0.92 (0.58-1.48) | 0.81 |
| *Shiga-like toxin-producing E. coli (O157)* | 10317 | 38/8677 (0.44) | 4/1640 (0.24) | ◆— | 0.56 (0.21-1.44) | 0.39 |
| *Shiga-like toxin-producing E. coli (non-O157)* | 10317 | 84/8677 (0.97) | 13/1640 (0.79) | ◆— | 0.82 (0.47-1.46) | 0.58 |
| *Shigella* species | 10317 | 97/8677 (1.12) | 9/1640 (0.55) | ◆— | 0.49 (0.26-0.93) | 0.033 |
| *Vibrio cholerae* | 10317 | 5/8677 (0.06) | 2/1640 (0.12) | ◆— | 2.12 (0.42-9.75) | 0.31 |
| *Vibrio* species | 10317 | 17/8677 (0.2) | 3/1640 (0.18) | ◆— | 0.93 (0.29-2.84) | >0.99 |
| *Yersinia enterocolitica* | 10317 | 58/8677 (0.67) | 13/1640 (0.79) | ◆— | 1.19 (0.66-2.15) | 0.52 |
| PARASITE |  |  |  |  |  |  |
| *Cryptosporidium* species | 10317 | 62/8677 (0.71) | 5/1640 (0.3) | ◆— | 0.42 (0.18-1.03) | 0.064 |
| *Cyclospora cayetanensis* | 10317 | 44/8677 (0.51) | 0/1640 (0) | ◆— | 0 (0-0.44) | 0.0007 |
| *Entamoeba histolytica* | 10317 | 4/8677 (0.05) | 0/1640 (0) | ◆— | 0 (0-5.3) | >0.99 |
| *Giardia lamblia* | 10317 | 100/8677 (1.15) | 11/1640 (0.67) | ◆— | 0.58 (0.32-1.05) | 0.09 |

To our knowledge, this is the first study to comprehensively evaluate the impact of COVID-19 pandemic-related social distancing measures on the gastrointestinal pathogen landscape. SIP orders and practice resulted in a notable decrease in human-to-human contact outside of households, with the closure of most workplaces, schools, and day cares, confinement of much of the population to their homes, as well as reduced travel (4). Although little data on food consumption patterns during SIP exist, we presume that the consumption of store-bought (including online purchases) and prepared restaurant foods (take-out and outdoor dining) largely persisted (5). In the absence of data to indicate significant changes in diet or food source, the altered gastrointestinal pathogen landscape that we observe is likely largely a result of decreased human-human contact and reduced travel. Although all the pathogens on the Biofire GI PCR panel are transmitted by the fecal-oral route, our findings suggest previously unappreciated differences in the transmission of these pathogens. Specifically, our data suggest that community spread of viral gastroenteritis, pathogenic *E. coli* (except for shiga toxin-producing *E. coli*), *Shigella*, and *Cyclospora* are more susceptible to changes associated with shelter-in-place than other gastrointestinal pathogens.

For adenovirus F40/41, astrovirus, norovirus, rotavirus, and sapovirus, transmission via person-to-person contact is well established (6). These pathogens often afflict daycares, nurseries, military barracks, and long-term care facilities, where personal contact is inevitable (6). It is conceivable that the closure of daycares and nurseries significantly reduced community transmission of these viruses, both among children and their close household contacts. Pathogenic *E. coli* and *Shigella*, on the other hand, are thought to be transmitted primarily through contaminated foods (7). Most outbreaks have been traced to specific foods, such as store-bought lettuce, flour, or ground beef (7). Our finding that these species (except for shiga toxin-producing *E. coli*) were detected significantly less frequently following SIP may reflect reduced travel-associated consumption of contaminated foods. In contrast, shiga toxin-producing strains of *E. coli* (O157 and non-O157), which are present in US-manufactured foods (8), were unaffected (7, 9). Similarly, incidence of the prototypical food-borne intestinal pathogens, *Campylobacter* and *Salmonella*, was unaffected, suggesting continuous production and consumption of contaminated foods.

We observed decreases in the rates of *Plesiomonas shigelloides, Cryptosporidium* species, *Entamoeba histolytica*, and *Giardia lamblia* rates following SIP. These differences were not statistically significant, and we attribute this to the relatively low frequency of these pathogens pre-SIP (see Figure 1). The decreased rates of these pathogens likely reflect reduced human-human contact, outdoor recreation, and travel, as contact with infected persons, exposure to contaminated recreational or drinking water, and travel to highly endemic areas are known risk factors for these pathogens (10-12). We observed a slight (12%) increase in the incidence of *Clostridium difficile* following SIP, which was not statistically significant (p = 0.23). Studies assessing the effect of COVID-19 on the incidence of *C. difficile* have focused on the hospital setting and reached differing conclusions. Ponce-Alonso *et al* (13) found that the rates of nosocomial *C. difficile* decreased by 70% in a tertiary care hospital in Spain during the COVID-19 pandemic, while Lewandowski *et al* (14) observed a four-fold increase in *C. difficile* infection at a hospital in Poland during the pandemic.

C*yclospora cayetanensis*, the etiologic agent of cyclosporiasis, is a parasite thought to be transmitted via contaminated food or water (15). Infected individuals shed unsporulated (non-infective) oocysts in their stool, which require 1-2 weeks in favorable environmental conditions to sporulate and become infective (15). It is therefore thought that person-to-person transmission is rare. Most outbreaks in the United States have been linked to imported fresh produce, such as raspberries, basil, and snow peas (16). Thus, the decrease in the incidence of C*yclospora cayetanensis* following SIP is most likely attributable to reduced travel, although reduced consumption of imported fresh produce as a result of disrupted food supply chains may have also contributed (17). Further epidemiological investigation will be required to test this hypothesis.

In summary, COVID-19 SIP measures represent a unique opportunity to reexamine our understanding of the transmission of infectious agents causing gastroenteritis. We identified shifts in the incidence of gastrointestinal pathogens following COVID-19 SIP that may shed new light on the transmission of these pathogens and provide opportunities to test novel strategies to disrupt transmissions of these pathogens. This study is limited in that it did not include patients with gastroenteritis that did not seek medical care and thus did not receive a FilmArray GI panel. The findings may be different if all patients with gastroenteritis were included. In addition, only cases dating back to January 2018 were included, precluding an in-depth analysis of annual season trends. The study is strengthened by a large sample size (>10,000 GI PCR assays), the unique circumstance of the California SIP, and the ability to precisely pinpoint the timing of SIP intervention. Additional studies are needed to better characterize the effect of COVID-19 and mitigation strategies on the transmission of gastrointestinal pathogens.

## Data Availability

Data may be requested by emailing the corresponding author.

